# Training AI to identify diagnostic criteria and treatment methods for advanced and delayed circadian sleep disorders in humans

**DOI:** 10.1101/2024.08.15.24312068

**Authors:** Lovemore Kunorozva, Ebele Okofor, Jacqueline M Lane

## Abstract

**Objectives:** The objective of this review was to evaluate the diagnosis and treatment of advanced and delayed sleep-wake phase disorders (ASWPD, DSWPD), both forms of circadian rhythm sleep-wake disorders (CRSWDs) using both human reviewers and ChatGTP to summarize relevant content.

**Methods:** PubMed-Medline, EbscoHost, and Web of Science were searched for peer reviewed articles. Original research articles published in English were searched for full-text studies. Studies that reported quantitative data on ASWPD and DSWPD diagnosing tools and treatment options in individuals of all ages were assessed. We assessed ChatGTP 4.0’s capacity to extract and summarize data from these studies and evaluated it alongside human reviewers.

**Results:** Our review of 49 articles on CRSWD from the past 20 years found that 91% focused on DSWPD. The most common diagnostic tools were the MEQ, MCQ, Pittsburgh Sleep Quality Index, and Epworth Sleepiness Scale, with primary methods including actigraphy and dim-light-melatonin-onset. Melatonin, often combined with light therapy and CBT, was the predominant treatment. ChatGPT-4.0 facilitated the review process with about 92% accuracy but required manual oversight for optimal results.

**Conclusions:** Our review identified key diagnostic tools, such as MEQ and MCQ surveys, and common treatment options for ASWPD and DSWPD, including melatonin, light therapy, and CBT. The findings underscore the need for comprehensive and individualized treatment approaches for CRSWDs, with a particular focus on expanding research on ASWPD and increasing data representation across age groups. While ChatGPT shows potential in streamlining data extraction for CRSWDs, it still requires human oversight to ensure accuracy.

## Introduction

Circadian rhythm sleep-wake disorder (CRSWD) is a condition that occurs when an individual’s internal sleep-wake schedule (i.e., biological clock) is out of sync with the natural light-dark cycles.^1,2^ This misalignment can occur because of the body’s inability to entrain to the 24-hour light-dark cycle or misalignment between one’s internal time and that of the external environment.^3^ CRSWDs often involve difficulty falling asleep, waking up too early or being unable to fall back to sleep once awake resulting in insomnia-like symptoms or excessive daytime sleepiness.^2^ Individuals who suffer from CRSWDs have shifted physiology and sleep/wake timing. Thus, CRSWDs can harm mental and physical health, as well as daily functioning.

The etiology of CRSWDs is complex and often involves both genetics and environmental factors.^4^ There are six main types of CRSWDs namely, advanced sleep-wake phase disorder (ASWPD),^2^ delayed sleep-wake phase disorder (DSWPD),^5^ irregular sleep-wake type disorder, non-24-hour sleep-wake disorder^5^ which is most often associated with blindness, jet-lag type, and shift-work disorder. Of these CRSWDs, ASWPD and DSWPD have demonstrated genetic susceptibility,^6^ with DSWPD accounting for most cases (3.3-4.6%)^7^ and being common among adolescents and youth while ASWPD is relatively rare affecting nearly 1% of individuals in middle to older age groups.

Patients with DSWPD tend to fall asleep very late and have delayed wake times compared with conventional times, which may result in chronic sleep insufficiency and daytime impairment due to differences between the “timing” of the world and the physiology.^7^ In contrast, patients with ASWPD tend to go to sleep and wake up early, often resulting in conflicts with evening social activities. In some cases, this syndrome may be caused by an inherited genetic disorder (i.e., it has a family tendency, FASWPD).^2^ Clinical diagnosis of CRSWD involves self-reported and objective methods such as dairies, questionnaires, sleep logs, dim light melatonin onset,^8^ and accelerometry tools such as actigraphy,^9^ and occasionally polysomnograms during a clinical visit.^10^ Also, treatment of CRSWD consists of sleep scheduling, behavioral modifications and education, timed melatonin administration and bright light exposure, and refraining from stimulating activities before bedtime.^2^

To date there is no consensus on the tools to clinically diagnose and treat CRSWD, leading to more misdiagnosis and treatment heterogeneity of ASWPD and DSWPD. Comparison of tools and treatment outcomes from individual studies is complicated by the use of inconsistent diagnostic and treatment criteria and the use of evening and morning chronotypes as proxies for DSWPD and ASWPD, respectively. The cause of ASWPD and DSWPD is unknown but may be related to genetics, underlying physical conditions, and a person’s behavior. This makes it difficult to diagnose and treat ASWPD and DSWPD, however, having a universally accepted approach to diagnosing and treating these CRSWDs would reduce opportunities for misdiagnosis. Additionally, this would enable a meta-analysis of ASWPD and DSWPD studies to be performed comparing diagnosis and treatment outcomes.

This systematic review aimed to provide comprehensive evidence of the tools used to diagnose and treat ASWPD and DSWPD. Furthermore, use this evidence to create a comprehensive chat engine review incorporating information from the various studies into a single tool, which can be used to improve diagnosis and treatment of ASWPD and DSWPD. This was achieved by using ChatGPT, a form of generative AI that lets users enter predefined prompts about ASWPD and DSWPD to receive human-like text, expediting data extraction and summarization. Furthermore, the dialogue format of ChatGPT makes it possible to test for accuracy, refine prompts and answer follow-up questions.

## Methodology

### Protocol

A protocol was developed according to guidelines outlined in the 2020 Preferred Reporting Items for Systematic Reviews and Meta-Analyses (PRISMA) statement.^11^ The research protocol was prospectively registered with the international Prospective Register Of Systematic Reviews (PROSPERO) in 2020 under registration number CRD566998. The PRISMA checklist is presented in the online supplemental file S1.

### Study selection and eligibility criteria

Eligibility criteria were established and agreed upon by all authors based on the concepts of population intervention and outcome. Studies that met the following criteria were considered eligible for inclusion in this systematic review:

1. Participants, male and female, clinically and self-reported to suffer from ASWPD and DSWPD.
2. All age groups, from children to older adults
3. Self-reported or clinically diagnosed ASWPD and/or DSWPD.
4. Treatment and/or behavioral therapies for individuals with ASWPD and/or DSWPD.
5. Before the search strategy implementation, it was agreed by all authors (LK, JL, EA) that journal articles with full-text original prospective and/or retrospective studies published in English would be included.

Exclusion criteria were set as studies:

1. Conducted with a heterogeneous sample (i.e., a mixed sample of ASWPD, DSWPD, and other sleep disorders) without reporting individual group findings separately.
2. Available as an abstract only (i.e., conference presentations), qualitative, discussion paper, commentary, or literature review.
3. Not available in English.

### Search strategy

Researchers systematically searched three electronic databases: PubMed-Medline, EBScoHost and Web of Science. Medical subject heading (MeSH) terms included: sleep-wake schedule disorder* OR sleep-wake schedule disorder* OR sleep-wake cycle disorder* OR Sleep-wake cycle disorder* OR circadian rhythm sleep disorder* OR advanced sleep phase syndrome* OR advanced sleep-phase syndrome* OR advanced sleep-wake phase disorder* OR delayed sleep phase syndrome* OR delayed sleep-wake phase disorder* OR delayed sleep-phase syndrome* OR familial advanced sleep-wake phase disorder* AND Diagnos* OR Examination* OR assessment* OR sign* OR treatment* AND Human AND relevant exclusions (see Supplementary File S2). A secondary search of the reference lists of included articles and hand searching in Google Scholar were performed. Further articles the authors were aware of relating to the topic were added to the search results. Duplicate articles were removed from the combined searches. Article screening and selection utilized the online tool CADIMA.^12^

**Figure 1:**
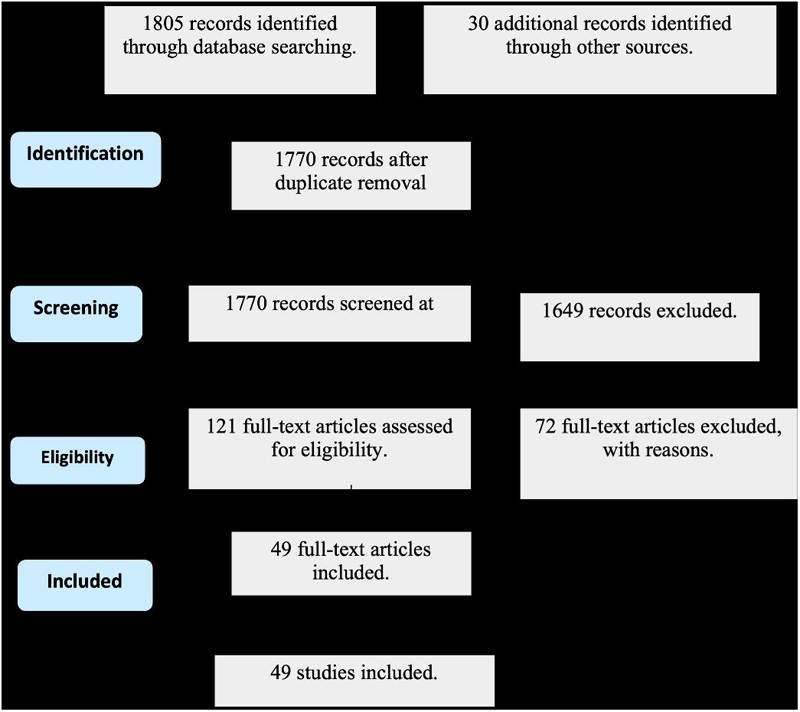
Preferred Reporting Items for Systematic Reviews and Meta-Analyses (PRISMA) flow diagram visualizing the selection process of identified, screened, and included articles following assessment of the eligibility criteria.

Two reviewers screened the articles independently (LK, EA). Full texts of articles were retrieved, and a second independent screening was undertaken by the same two reviewers. Any conflicts were resolved through discussion and consensus between reviewers. A third reviewer (JL) served as the tie-breaking vote.

### Data extraction

To implement artificial intelligence for data extraction and summarization, ChatGTP was tested for accuracy and to refine the prompts. To accomplish this, data from two publications was manually extracted. A list of prompts was generated to extract data using ChatGTP from the test publications. Each publication was entered into ChatGTP and a list of prompts was used to extract data testing for accuracy. Prompts were then iteratively revised until ChatGTP data extraction prompts and accuracy had been improved. Following this process, data from the remaining 47 articles was extracted. Extracted data included: participant details (number, age, gender), study design, sample size, diagnosis (ASWPD or DSWPD), treatment or intervention, population group (children: 0-12 years old, youth/adolescent: 13-17 years old, or adult: 18-100 years old), and diagnosis tool (polysomnography, actigraphy or sleep log or combination thereof).

### Quality Assessment and risk of bias

Studies were reviewed for the quality assessment and risk of bias using a modified Downs and Black tool^13^ (see Supplementary S3). This was done by three reviewers (LK, EO, JL) independently scoring the articles and then discussing differences to reach an agreed final score for each article. The same reviewers determined the level of evidence using the Oxford Centre for Evidence-Based Medicine (OCEBM, 2009).^14^ The articles fell into two main categories: Observational studies of the diagnoses of ASWPD or DSWPD; or Interventional studies where the treatment outcome was determined in response to the intervention, with or without control groups. The OCEBM level of evidence was graded using the criteria for a Symptom Prevalence Study for the observational studies based on the degree of follow up for prospective studies as level 1b for good follow-up and level 3b for non-consecutive cohort studies, and level 2b for retrospective studies. The intervention studies were graded using the Therapy/Prevention studies criteria of level 1b for randomized control trials (RCTs) with narrow confidence intervals and level 2b for the non-RCT studies.

## Results

### Literature search

A total of 1770 studies were identified in the core collection databases and additional sources for screening, after duplicates were removed. Figure 1 (PRISMA flow diagram) summarizes the study selection process and reasons for excluding studies. The full texts of 121 articles were assessed for eligibility, 72 were excluded and 49 were included.

#### Quality assessment

The quality assessment of the 49 studies using the modified Downs and Black score^13^ is shown in Supplementary File S3. The quality assessment showed that two studies^15,16^ (4%) was scored as fair (score 6-8), nine studies^17–23^ (18%) were scored ‘good’ (score 9-10), 39 studies (78%) were scored ‘excellent’ (score 11-13). Using the OCEBM scale for observational studies,^14^ 22 studies (44.9 %) were rated as 1b level of evidence, 7 studies^7,24–29^ (14.3 %) were rated as 2b level of evidence, and 20 studies (40.8 %) were rated as 3b level of evidence (Supplementary File S3). There were only nine interventional studies included (18.4 %) of which five were RCTs with narrow confidence intervals^16,25,30–34^ rated as level 1b and the other four studies were non-RCT^24,25,29^ rated as level 2b.

#### Characteristics of included studies

The characteristics of the 49 studies are presented in Table 1. The age range of participants included in this review was 7 to 65 years. The 49 studies had a total of 81,119 participants (range: 3 to 50,054), including 32,575 (40.16%) males, 48,509 (59.8%) females, and 35 (0.04%) unspecified sex. Studies were conducted across 10 different countries and four different continents. Fifty percent of the studies in this review were conducted in Australia (N=12) and Japan (N=12). Eighteen studies (37%) had a focus on diagnoses only, 17 studies (35%) focused on treatment only, and 14 studies (29%) focused on both diagnoses and treatment.

### Diagnosis of ASWPD and DSWPD

Of the 49 studies, two studies (4.1%) investigated ASWPD, two studies (4.1%) investigated both ASWPD and DSWPD^35^ and 45 studies (91.2%) investigated DSWPD (Figure 1A). Thirty-three studies focused on the diagnosis of individuals with ASWPD/DSWPD. Of the 33 studies (Figure 1D), nine studies (26.4%) used a combination of actigraphy, DLMO and validated surveys reviewed by a clinician, eight studies (23.5%) used a combination of DLMO, actigraphy and sleep logs or diaries, seven studies (20.6%) used sleep logs or validated questionnaires, four studies (11.8%) used sleep logs and validated questionnaires reviewed by a clinician, three studies (8.8%) used a combination of PSG, DLMO, actigraphy, sleep logs and validated surveys, with one of these studies reviewed by a clinician. Lastly, in two studies (5.9%) participants were diagnosed using PSG^26,36^ only and in one study (2.9%)^35^ participants were diagnosed based on self-reporting only (Figure 1D).

Of the validated questionnaires used the Morningness-Eveningness Questionnaire and Munich Chronotype Questionnaire (n=20, 35.7%), Pittsburgh Sleep Quality Index (n=10, 17.9%) and the Epworth Sleepiness Scale (n=9, 16.1%) were the most frequently used (Figure 1C). The least used questionnaire included Karolinska^29,33,37^ (n=3, 5.4%) Insomnia Severity Index^30,38^ (n=2, 3.6%), DSWPD-STQ^22,24,39^ (n=3, 5.4%), Hospital Depression and Anxiety Scale^33,38^ (n=2, 3.6%) and the Depression, Anxiety and Stress Scale^39,40^ (n=2, 3.6%, Figure 1C)

### Treatment options for ASWPD and DSWPD

Of the 49 studies, 17 studies (34.7%) investigated treatment options for individuals with ASWPD/DSWPD. The treatment options are presented in Figure 1B. Melatonin (n=14, 37.8%), light therapy (n=11, 29.7%), and Cognitive and behavioral therapy (CBT, n=8, 21.6%) were the most used treatment options. More specifically, six studies (35.3%) used melatonin only, three studies (17.6%) used a combination of melatonin, light therapy, and CBT, two studies (11.8%) used melatonin, chrono- and light therapy, one study (5.9%) used melatonin and light therapy, three studies (17.6%) used CBT and light therapy, with the remaining two studies using ramelteon^15^ (5.9%) and CBT (5.9%) only (Figure 1B).

**Figure 2:**
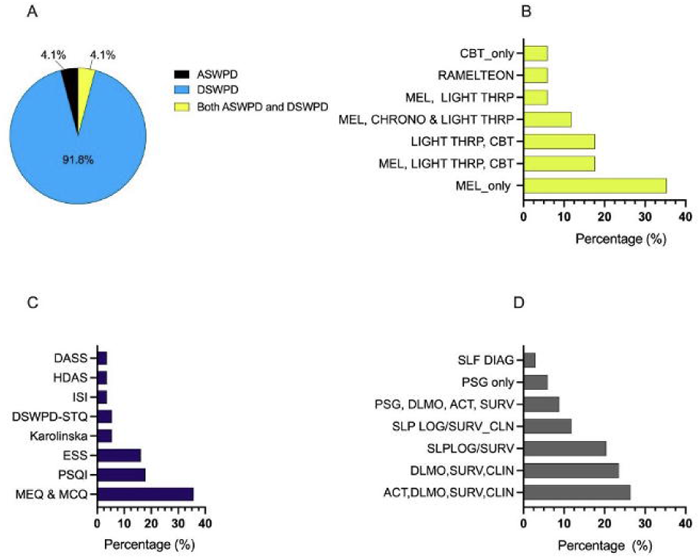
Figure showing the percentage of (A) advanced and delayed phase sleep-wake disorder, (B) treatment options, (C) validated surveys and (D) diagnostic approaches used in the various studies reviewed. Note: ASWPD-advanced sleep-wake phase disorder, DSWPD – delayed sleep-wake phase disorder, CBT - chronic and cognitive behavioral therapy, MEL - melatonin, CHRONO THRP – chrono-therapy, LIGHT THRP-light therapy, DASS – Depression, anxiety and stress scale, HDAS – hospital depression and anxiety scale, ESS – Epworth Sleepiness Scale, PSQI – Pittsburgh Sleep Quality Index, MEQ – Morningness-Eveningness personality Questionnaire, MCQ – Munich Chronotype Questionnaire, ACT – actigraphy, DLMO – dim light melatonin onset, CLIN – clinician reviewed, SURV – validated survey, DSWPD-STQ – delayed sleep-wake phase disorder sleep timing questionnaire, SLP LOG – sleep log, PSG – polysomnography, SLF_DAG - self-diagnosis.

### CHATGTP and CRSWD data extraction

The initial data extraction from a subset of articles (n=4) achieved an accuracy and relevance rate of 80%. This prompted revisions to refine the extraction criteria, which subsequently enhanced the precision and relevance of the extracted information. By optimizing the extraction parameters, we reduced irrelevant wording and increased the specificity of the queries, ensuring a closer match to the original article. Following these refinements, blind extraction conducted independently by two reviewers showed consistent reproducibility across all four studies during the second stage of review. The accuracy of the extracted data, including critical details such as study population demographics (age and gender), was meticulously verified against the original articles. This verification process confirmed a 92% accuracy rate in the extracted data, ensuring both completeness and reliability. Standardized extraction parameters proved effective across a diverse range of studies, facilitating consistency in data extraction. This standardization enabled automation of the 49 studies, significantly expediting the data extraction process and enhancing overall efficiency.

## Discussion

This systematic review aimed to summarize the diagnostic themes and treatment options for ASWPD and DSWPD as reported in the literature, to assess consistency and inform future research. Additionally, we evaluated ChatGTP’s effectiveness in expediting data extraction and summarization process for CRSWD from studies. Specifically, it sought to present a comprehensive overview of the current evidence regarding the diagnosis and treatment of ASWPD and DSWPD across diverse populations and age groups. DSWPD emerged as the most frequently reported CRSWD in both diagnostic and treatment contexts, with only two studies^9,35^ addressing ASWPD diagnoses and treatment. The discussion within this systematic review centers on three key areas of interest: 1) diagnostic strategies; 2) treatment modalities available for individuals affected by both ASWPD and DSWPD and 3) an evaluation of ChatGPT’s capability to expedite and accurately summarize study criteria.

### Diagnostic approaches for ASWPD and DSWPD

Forty-five of the reviewed studies focused on DSWPD, while two addressed ASWPD,^9,41^ and an additional two studies^35^ covered both ASWPD and DSWPD. The findings from this review revealed a notably higher prevalence of DSWPD compared to ASWPD within the studied population, aligning with existing research indicating that DSWPD is more prevalent.^7,38,42^ This observation is likely due to most participants being adolescents and young adults (16-35 years old), who have shown a prevalence of mostly environmentally induced DSWPD ranging from 3.3% to 4.6%.^42^ Young adults and adolescents deal with various social and lifestyle pressures, such as academic demands and extracurricular activities and have relatively lose social constraints, which frequently disrupt their sleep patterns. Factors like increased electronic device usage, irregular sleep schedules, and late-night engagements such as studying or socializing further contribute to delays in sleep onset.

Moreover, during adolescence and early adulthood, a delay in circadian phase preference, influenced in part by genetic factors, has been identified.^42^ This delay leads to sleep onset insomnia, difficulty waking up at the desired time, and associated daytime dysfunction. Additionally, heightened exposure to light in the evening can suppress melatonin production, further delaying sleep onset. Consequently, a combination of biological, social, and environmental factors renders adolescents and young adults particularly susceptible to experiencing DSWPD. Understanding these multifaceted influences is crucial for developing tailored prevention and management strategies for this population group.

Of the 49 studies included in this review, 33 studies centered on diagnosing individuals with ASWPD/DSWPD utilized a combination of actigraphy, sleep logs or diaries, DLMO, and validated surveys reviewed by a clinician. These diagnosing approaches offer a comprehensive assessment of various aspects related to circadian rhythm and sleep patterns. For example, DLMO provides a direct measure of an individual’s circadian phase, while actigraphy allows for objective monitoring of sleep-wake patterns over an extended period. Sleep logs or diaries provide subjective data on sleep habits and quality, complementing the objective measures obtained from DLMO and actigraphy. Utilization of these approaches seeks to balance patient and clinician burden and cost.

The literature on the diagnosis and treatment of CRSWDs generally aligns with the DSM diagnostic criteria in recognizing the importance of assessing sleep patterns, circadian rhythms, and the impact on daily functioning. Both sources emphasize the necessity of a thorough clinical evaluation, including sleep history and objective sleep measurements. However, the literature often highlights a broader range of treatment options, including chronotherapy^32^ and light therapy,^20,37^ which are not prominently featured in the DSM guidelines. Additionally, the literature underscores the role of genetic factors more explicitly than the DSM, indicating a potential area for further integration. Moving forward, establishing a consensus between the diagnostic criteria and treatment options outlined in the literature and those highlighted in the DSM is essential to enhance the well-being of individuals suffering from CRSWDs.

The Morningness-Eveningness Questionnaire^43^ and Munich Chronotype Questionnaires, along with the Pittsburgh Sleep Quality Index^44^ and the Epworth Sleepiness Scale,^45^ were the most frequently utilized questionnaires in diagnosing ASWPD and DSWPD. Conversely, the Depression Anxiety Stress Scales,^46^ Hypersomnia Diagnostic Assessment Scale, Insomnia Severity Index, Karolinska Sleepiness Scale, and DSWPD Sleep Timing Questionnaire were less commonly employed. Depression Anxiety Stress Scales, Hypersomnia Diagnostic Assessment Scale, Insomnia Severity Index, Karolinska Sleepiness Scale, and DSWPD Sleep Timing Questionnaire are not frequently used in the diagnosis of CRSWDs because they primarily assess other aspects of sleep such as depression, anxiety, stress, hypersomnia, insomnia severity, and sleepiness levels rather than directly evaluating circadian rhythm disturbances.

The former set of tools, including the MEQ, ESS, PSQI, and MCQ, are favored for their ability to comprehensively assess an individual’s circadian rhythm and sleep patterns, essential in determining the severity of ASWPD and DSWPD. Finally, having these assessments reviewed by a clinician ensures thorough interpretation and integration of the data, leading to a more accurate diagnosis and personalized treatment approach tailored to the individual’s needs.

### Treatment options for ASWPD and DSWPD

Seventeen studies investigated treatment options for individuals with ASWPD/DSWPD. The significant proportion of studies investigating treatment options for individuals with ASWPD and DSWPD highlights the growing interest in developing effective interventions to manage these CRSWD. Among the various treatment modalities explored, melatonin emerged as the most frequently utilized option, reflecting its role in regulating the sleep-wake cycle and its efficacy in addressing circadian rhythm disruptions. Moreover, the substantial utilization of light therapy underscores its importance in synchronizing the circadian rhythm with the external light-dark cycle, particularly beneficial for individuals experiencing delayed sleep-wake phase. Light therapy^47^ offers a non-pharmacological approach to resetting the circadian clock and promoting earlier sleep onset and wake-up times. CBT^16^ also garnered attention as a treatment option, highlighting the recognition of the psychological and behavioral factors that influence sleep patterns. CBT interventions aim to address maladaptive sleep behaviors and thought patterns, promoting healthy sleep habits and enhancing overall sleep quality.

The combination of melatonin, light therapy, and CBT in some studies reflects the adoption of a multi-modal approach to treatment, leveraging the synergistic effects of these interventions to address various aspects of circadian-aligned ASWPD and DSWPD disturbances. Additionally, using other pharmacological agents such as ramelteon underscores the ongoing exploration of novel treatment options for ASWPD and DSWPD and the need for further pharmaceuticals aimed at treatment of circadian disruption. Overall, the diverse array of treatment modalities investigated underscores the complexity of CRSWD and the need for individualized approaches to management. Further research is warranted to elucidate the comparative efficacy and long-term outcomes of these interventions, ultimately improving the quality of care for individuals affected by ASWPD and DSWPD.

The ChatGTP findings highlight the effectiveness of refining data extraction criteria, resulting in a significant increase in accuracy and relevance of the extracted data, with accuracy improving from 80% to over 90%. The use of standardized extraction parameters not only ensured consistency across various studies but also allowed for successful automation of the extraction process, thereby enhancing efficiency. The reproducibility of data extracted by independent reviewers further validated the robustness of the revised extraction methodology. Thus, using AI for performing literature reviews in the CRSWD space significantly enhances efficiency, comprehensiveness, and accuracy. It can swiftly scan and analyze extensive literature, automating searches across various articles, which greatly reduces the time required for traditional reviews. It integrates both structured (Text) and unstructured (Tables and Figures) data, ensuring a comprehensive review that incorporates multiple data streams

By applying consistent criteria for evaluating literature, ChatGPT minimizes human errors and biases, leading to more reliable and accurate systematic reviews. Its customizable searches and ability to handle large data volumes make it ideal for extensive reviews. Moreover, ChatGPT’s cost-effectiveness and scalability save resources, making the literature review process in the CRSWD space more accessible and efficient for researchers, ultimately leading to higher-quality research outcomes.

### Limitations

The lack of demographic data concerning race, ethnicity, and income levels in some reviewed studies poses a limitation to the generalizability of these findings. The gender bias and predominant focus on adults in the reviewed literature may inadvertently neglect the specific needs of other demographic groups. Furthermore, the reliance on self-reported surveys in certain studies introduces the potential for bias in diagnosis. These factors underscore the importance of considering diverse demographic factors and employing objective assessment methods to ensure the comprehensive understanding and accurate diagnosis of circadian rhythm sleep disorders. A limitation of these findings is that the reliance on automated data extraction and standardized parameters may overlook context-specific nuances, potentially leading to missed or misinterpreted information that requires manual review to fully capture. This review assessed only diagnostic and treatment options published in English.

We may have excluded some studies which did not meet strict inclusion criteria, potentially omitting important evidence. based on our inclusion criteria. We faced challenges in combining data from different studies due to differences in study design or measurement tools. Challenges were also encountered in combining data from different studies due to variations in study design and measurement tools, which prohibited our ability to conduct a meaningful meta-analysis or draw cohesive conclusions on treatment and diagnostic criteria. The reviewed studies may not be generalizable to other populations or settings, such as those in Africa and developing countries, where no studies on CRSWD were found. Consequently, the review findings might be limited primarily to individuals of European descent.

## Conclusion

This systematic review comprehensively summarized and identified various diagnostic criteria and treatment options currently employed in different clinical and study settings. Additionally, it evaluated ChatGTP’s effectiveness in data extraction and summarization for ASWPD and DSWPD. DSWPD emerged as the predominant focus for many studies, reflecting its higher prevalence compared to ASWPD. Diagnostic strategies primarily involved a combination of objective measures such as actigraphy and DLMO, complemented by subjective assessments like validated questionnaires.

Treatment modalities predominantly included melatonin, light therapy, and CBT, with some studies exploring multi-modal approaches. These findings underscore the complexity of CRSWDs and the need for individualized, multi-faceted treatment approaches to effectively manage ASWPD and DSWPD. It also highlights the need to establish a consensus between the diagnostic criteria and treatment options outlined in the literature and those highlighted in the DSM to enhance the well-being of individuals suffering from CRSWDs. Further research is warranted to refine diagnostic techniques and explore additional therapeutic interventions to improve outcomes for affected individuals. Lastly, while ChatGPT is an effective tool for expediting data extraction and summarization, human oversight is necessary to ensure improved accuracy and relevance.

## Supporting information

PRISMA CHECKLIST

## Data Availability

All data produced in the present work are contained in the manuscript

## References

1. Abbott, S. M., Malkani, R. G. & Zee, P. C. Circadian disruption and human health: A bidirectional relationship. Eur. J. Neurosci. 51, 567–583 (2020).

2. Auger, R. R. et al. Clinical Practice Guideline for the Treatment of Intrinsic Circadian Rhythm Sleep-Wake Disorders: Advanced Sleep-Wake Phase Disorder (ASWPD), Delayed Sleep-Wake Phase Disorder (DSWPD), Non-24-Hour Sleep-Wake Rhythm Disorder (N24SWD), and Irregular Sleep-Wake Rhythm Disorder (ISWRD). An Update for 2015: An American Academy of Sleep Medicine Clinical Practice Guideline. J. Clin. Sleep Med. 11, 1199–1236 (2015).

3. Mistlberger, R. E. & Skene, D. J. Nonphotic entrainment in humans? J. Biol. Rhythms 20, 339–352 (2005).

4. Ashbrook, L. H., Krystal, A. D., Fu, Y.-H. & Ptáček, L. J. Genetics of the human circadian clock and sleep homeostat. Neuropsychopharmacology 45, 45–54 (2020).

5. Abbott, S. M., Choi, J., Wilson, J. & Zee, P. C. Melanopsin-dependent phototransduction is impaired in delayed sleep–wake phase disorder and sighted non–24-hour sleep–wake rhythm disorder. Sleep 44, zsaa184 (2021).

6. Liu, W. Mini-review: Genetics of common types of sleep disorders. in 020015 (Chongqing City, China, 2019). doi:10.1063/1.5085528.

7. Sivertsen, B., Harvey, A. G., Gradisar, M., Pallesen, S. & Hysing, M. Delayed sleep–wake phase disorder in young adults: prevalence and correlates from a national survey of Norwegian university students. Sleep Med. 77, 184–191 (2021).

8. Keijzer, H., Spruyt, K., Smits, M. G., de Geest, A. & Curfs, L. M. G. Can dim light melatonin onset be predicted by the timing of sleep in patients with possible circadian sleep-wake rhythm disorders? Biol. Rhythm Res. 48, 557–566 (2017).

9. Björkqvist, J. et al. Advanced sleep–wake rhythm in adults born prematurely: confirmation by actigraphy-based assessment in the Helsinki Study of Very Low Birth Weight Adults. Sleep Med. 15, 1101–1106 (2014).

10. Roh, H. W. et al. Associations of actigraphy derived rest activity patterns and circadian phase with clinical symptoms and polysomnographic parameters in chronic insomnia disorders. Sci. Rep. 12, 4895 (2022).

11. Liberati, A. et al. The PRISMA statement for reporting systematic reviews and meta-analyses of studies that evaluate health care interventions: explanation and elaboration. J. Clin. Epidemiol. 62, e1–e34 (2009).

12. Kohl, C. et al. Online tools supporting the conduct and reporting of systematic reviews and systematic maps: a case study on CADIMA and review of existing tools. Environ. Evid. 7, 8 (2018).

13. Downs, S. H. & Black, N. The feasibility of creating a checklist for the assessment of the methodological quality both of randomised and non-randomised studies of health care interventions. J. Epidemiol. Community Health 52, 377–384 (1998).

14. The 2011 Oxford CEBM Levels of Evidence: Introductory Document. 1–3 (2011).

15. Izuhara, M., Kawano, K., Otsuki, K., Hashioka, S. & Inagaki, M. Prompt improvement of difficulty with sleep initiation and waking up in the morning and daytime somnolence by combination therapy of suvorexant and ramelteon in delayed sleep-wake phase disorder: a case series of three patients. Sleep Med. 80, 100–104 (2021).

16. Jansson-Fröjmark, M., Danielsson, K., Markström, A. & Broman, J.-E. Developing a cognitive behavioral therapy manual for delayed sleep–wake phase disorder. Cogn. Behav. Ther. 45, 518–532 (2016).

17. Burgess, H. J., Park, M., Wyatt, J. K. & Fogg, L. F. Home dim light melatonin onsets with measures of compliance in delayed sleep phase disorder. J. Sleep Res. 25, 314–317 (2016).

18. Carriere, C., Coste, O., Meiffred-Drouet, M., Barat, P. & Thibault, H. Sleep disorders in obese children are not limited to obstructive sleep apnoea syndrome. Acta Paediatr. 107, 658–665 (2018).

19. Ebisawa, T. et al. Association of structural polymorphisms in the human *period3* gene with delayed sleep phase syndrome. EMBO Rep. 2, 342–346 (2001).

20. Joo, E. Y. et al. Timing of light exposure and activity in adults with delayed sleep-wake phase disorder. Sleep Med. 32, 259–265 (2017).

21. Lovato, N., Gradisar, M., Short, M., Dohnt, H. & Micic, G. Delayed Sleep Phase Disorder in an Australian School-Based Sample of Adolescents. J. Clin. Sleep Med. 09, 939–944 (2013).

22. Micic, G. et al. The endogenous circadian temperature period length (tau) in delayed sleep phase disorder compared to good sleepers. J. Sleep Res. 22, 617–624 (2013).

23. Schubert, J. R. & Coles, M. E. Obsessive-Compulsive Symptoms and Characteristics in Individuals With Delayed Sleep Phase Disorder. J. Nerv. Ment. Dis. 201, 877–884 (2013).

24. Micic, G. et al. Readiness to change and commitment as predictors of therapy compliance in adolescents with Delayed Sleep-Wake Phase Disorder. Sleep Med. 55, 48–55 (2019).

25. Richardson, C. et al. Cognitive performance in adolescents with Delayed Sleep-Wake Phase Disorder: Treatment effects and a comparison with good sleepers. J. Adolesc. 65, 72–84 (2018).

26. Rubens, S. L., Patrick, K. E., Williamson, A. A., Moore, M. & Mindell, J. A. Individual and socio-demographic factors related to presenting problem and diagnostic impressions at a pediatric sleep clinic. Sleep Med. 25, 67–72 (2016).

27. Sivertsen, B., Harvey, A. G., Pallesen, S. & Hysing, M. Mental health problems in adolescents with delayed sleep phase: results from a large population-based study in Norway. J. Sleep Res. 24, 11–18 (2015).

28. Sivertsen, B. et al. Delayed sleep phase syndrome in adolescents: prevalence and correlates in a large population based study. BMC Public Health 13, 1163 (2013).

29. Solheim, B. et al. Sleep structure and awakening threshold in delayed sleep-wake phase disorder patients compared to healthy sleepers. Sleep Med. 46, 61–68 (2018).

30. Sletten, T. L. et al. Efficacy of melatonin with behavioural sleep-wake scheduling for delayed sleep-wake phase disorder: A double-blind, randomised clinical trial. PLOS Med. 15, e1002587 (2018).

31. Takaesu, Y. et al. Circadian rhythm sleep-wake disorders as predictors for bipolar disorder in patients with remitted mood disorders. J. Affect. Disord. 220, 57–61 (2017).

32. Van Andel, E., Bijlenga, D., Vogel, S. W. N., Beekman, A. T. F. & Kooij, J. J. S. Effects of chronotherapy on circadian rhythm and ADHD symptoms in adults with attention-deficit/hyperactivity disorder and delayed sleep phase syndrome: a randomized clinical trial. Chronobiol. Int. 38, 260–269 (2021).

33. Wilhelmsen-Langeland, A. et al. The Personality Profile of Young Adults With Delayed Sleep Phase Disorder. Behav. Sleep. Med. 12, 481–492 (2014).

34. Saxvig, I. W., Pallesen, S., Wilhelmsen-Langeland, A., Molde, H. & Bjorvatn, B. Prevalence and correlates of delayed sleep phase in high school students. Sleep Med. 13, 193–199 (2012).

35. Paine, S.-J., Fink, J., Gander, P. H. & Warman, G. R. Identifying advanced and delayed sleep phase disorders in the general population: A national survey of New Zealand adults. Chronobiol. Int. 31, 627–636 (2014).

36. Saxvig, I. W. et al. Objective measures of sleep and dim light melatonin onset in adolescents and young adults with delayed sleep phase disorder compared to healthy controls. J. Sleep Res. 22, 365–372 (2013).

37. Moderie, C., Van Der Maren, S. & Dumont, M. Circadian phase, dynamics of subjective sleepiness and sensitivity to blue light in young adults complaining of a delayed sleep schedule. Sleep Med. 34, 148–155 (2017).

38. Danielsson, K., Markström, A., Broman, J.-E., Von Knorring, L. & Jansson-Fröjmark, M. Delayed sleep phase disorder in a Swedish cohort of adolescents and young adults: Prevalence and associated factors. Chronobiol. Int. 33, 1331–1339 (2016).

39. Lovato, N. et al. Can the circadian phase be estimated from self-reported sleep timing in patients with Delayed Sleep Wake Phase Disorder to guide timing of chronobiologic treatment? Chronobiol. Int. 33, 1376–1390 (2016).

40. Micic, G., Lovato, N., Gradisar, M. & Lack, L. C. Personality differences in patients with delayed sleep–wake phase disorder and non-24-h sleep–wake rhythm disorder relative to healthy sleepers. Sleep Med. 30, 128–135 (2017).

41. Hibbs, A. M. et al. Advanced Sleep Phase in Adolescents Born Preterm. Behav. Sleep. Med. 12, 412–424 (2014).

42. Tomishima, S., Komada, Y., Tanioka, K., Okajima, I. & Inoue, Y. Prevalence and Factors Associated With the Risk of Delayed Sleep-Wake Phase Disorder in Japanese Youth. Front. Psychiatry 13, 878042 (2022).

43. Horne, J. A. & Ostberg, O. A self assessment questionnaire to determine Morningness Eveningness in human circadian rhythms. Int. J. Chronobiol. 4, 97–110 (1976).

44. Buysse, D. J., Reynolds, C. F., Monk, T. H., Berman, S. R. & Kupfer, D. J. The Pittsburgh sleep quality index: A new instrument for psychiatric practice and research. Psychiatry Res. 28, 193–213 (1989).

45. Gelaye, B. et al. Construct Validity and Factor Structure of the Pittsburgh Sleep Quality Index and Epworth Sleepiness Scale in a Multi-National Study of African, South East Asian and South American College Students. PLoS ONE 9, e116383 (2014).

46. Kroenke, K., Spitzer, R. L., Williams, J. B. W. & Löwe, B. An Ultra-Brief Screening Scale for Anxiety and Depression: The PHQ–4. Psychosomatics 50, 613–621 (2009).

47. Baiardi, S. et al. Chronobiology, sleep-related risk factors and light therapy in perinatal depression: the “Life-ON” project. BMC Psychiatry 16, 374 (2016).

